# Characterizing the Burden of Scrub Typhus in Nepalese Children using Representative School-Based Cross-Sectional Serosurveys

**DOI:** 10.64898/2026.08.28.26361601

**Authors:** Shiva Ram Naga, Sabin Bikram Shahi, Sarita Gosain, Mamata Maharjan, Nishan Katuwal, Ezra Morrison, Jason Andrews, Rajeev Shrestha, Dipesh Tamrakar, Kristen Aiemjoy

## Abstract

**Background:** Scrub typhus, an acute bacterial infection caused by *Orientia tsutsugamushi*, is an important, under-recognized etiology of febrile illness in South Asia. Accurate surveillance for scrub typhus in Nepal is challenging due to non-specific symptoms and limited diagnostic tools.

**Methods:** We conducted a representative school-based cross-sectional serosurvey in Kavrepalanchok and Dolakha, Nepal between November 2021 and April 2022 using two-stage sampling, we randomly selected 13 public schools and then up to 100 children aged 4-18 years per school. We collected capillary blood samples and tested for IgG responses to *Orientia tsutsugamushi*-derived recombinant 56-kDa antigen using commercially available ELISA kits. We estimated seroincidence rates using previously-published models of antibody decay dynamics from confirmed scrub typhus cases. We compared seroincidence to seroprevalence estimates using cutoffs derived from Gaussian finite mixture models applied to the study population.

**Results:** We enrolled a total of 827 children (participation rate: 94.8%). The median age was 10 years (IQR: 8–13), and 53.08% of the participants were female. The overall seroincidence rate was 6.3 new infections per 1000 person-years (95% CI: 4.7- 8.4), and the rate was slightly higher in peri-urban Kavrepalanchok (7.3; 95% CI: 4.9 - 10.3) compared with rural Dolakha (5.3; 95% CI: 3.3 - 8.5). Seroincidence increased with age, from 3.6 new infections per 1000 person-years (95% CI: 1.4- 9.7) among children aged 4- 7 years to 9.4 (95% CI: 6.2-14.3) among children aged 14-18 years. Seroincidence was higher in females (8.3; 95% CI: 5.9 -11.8) than in males (4.1; 95% CI: 2.4 - 6.9) (p=0.02). The overall seroprevalence was 6.5% (95% CI: 4.9–8.4) and followed a similar age and geographic trend to the seroincidence rate.

**Discussion:** Our findings reveal a substantial burden of pediatric scrub typhus in the Kavrepalanchok and Dolakha districts of Nepal. Seroincidence increased with age and was higher among females. School-based serosurveys offer an efficient sampling frame to rapidly assess population-level scrub typhus transmission intensity among children and adolescents, though may not represent out-of-school populations.

## BACKGROUND

Scrub typhus, a severe bacterial infection caused by *Orientia tsutsugamushi*, is an underrecognized cause of morbidity and mortality in the Asia Pacific region (1–3). Infection occurs when trombiculid mite larva (chiggers) feed on host skin cells. The mite larvae are so small that the bite is typically painless and unrecognized(4). Scrub typhus typically presents with nonspecific symptoms such as fever and headache. If untreated, it can progress to severe complications including pneumonia, myocarditis, and meningoencephalitis, which can be fatal(5).

While scrub typhus is endemic in Nepal, the exact burden is not well-defined (6). An early scrub typhus serosurvey conducted in 1981 indicated a seroprevalence of 10% among healthy adults aged 17-50 year-old males(7). Before 2014, there was no clear evidence of significant outbreaks or fatalities from scrub typhus in Nepal, and government investigations and surveillance were limited(6). The disease was recognized as a significant etiology of febrile illness after the 2015 earthquake when several outbreaks of scrub typhus were reported from different parts of Nepal (8,9). Scrub typhus is endemic in Nepal’s neighboring countries, including India, China and Bhutan(1,9–11).

There are no licensed vaccines for scrub typhus, nor are there organized efforts for vector control (12,13). Exposure to mites usually occurs during contact with the ground and vegetation; therefore, people living in rural areas, especially agricultural workers, are at heightened risk(14). However, exposure also occurs in urban and peri-urban settings(15). A substantial portion of scrub typhus infections remain undetected because of the lack of diagnostic tools and limited awareness, resulting in underreporting and an underestimation of the disease’s overall impact(16).

Serological surveillance can augment clinical surveillance to characterize scrub typhus epidemiology, including prevalence, spatial distribution, and identifying at-risk populations(17). Two measures of burden are typically reported from serosurveillance studies: seroprevalence, which is the percentage of individuals seropositive at a given time point, and seroincidence (also known as force of infection), which is the rate at which new infections occur in a population. Seroprevalence is affected by the age distribution of the population and the rate at which antibodies decay over time. Our group previously conducted population-based serosurveillance in Kathmandu and Kavrepalanchok districts and developed methods to calculate seroincidence from cross-sectional serosurveys, accounting for antibody waning (17). Using antibody dynamics data characterized from clinical cohorts of confirmed scrub typhus cases, we estimated that scrub typhus IgG antibodies decay with a half-life of approximately 2 years. Applying these antibody kinetics parameters to cross-sectional data, we estimated that the annual seroincidence rates in these districts ranged from 5 to 15 per 1000 person-years(17).

While population-based samples are considered the ideal sampling frame for generating representative estimates of infection burden, schools are increasingly recognized as an alternative population that can be used to efficiently characterize infection burden among children (18–20). Schools are logistically easier and faster to sample and therefore are valuable for rapid serosurveillance. Children also represent an important population for scrub typhus surveillance as they may have different exposure patterns than adults(21,22). However, it is unknown if school-based sampling introduces bias relative to representative population-based sampling frames.

To address this methodological gap, we conducted a cross-sectional serosurvey with two objectives: first, to estimate the seroincidence and seroprevalence of scrub typhus among school-aged children in Kavrepalanchok and Dolakha districts, Nepal; and second, to evaluate whether school-based sampling provides comparable estimates to population-based sampling for scrub typhus serosurveillance. Kavrepalanchok is a peri-urban district where our previous studies demonstrated moderate to high scrub typhus burden, whereas Dolakha is a rural district with less well-characterized transmission patterns.

## METHODS

### Study Design and Population

We conducted a school-based cross-sectional enteric fever serosurvey among children enrolled in public and private schools in Kavrepalanchok and Dolakha districts, Nepal between November 2021 and April 2022. In this nested substudy, we tested residual samples collected from children enrolled in public schools for scrub typhus.

Kavrepalanchok is a peri-urban district that borders the capital, Kathmandu with a population of approximately 364,039, while Dolakha is a rural, mountainous district with a population of approximately 172,767(23).

In Kavrepalanchok, we used stratified sampling to select one public school from each of four municipalities (Dhulikhel, Banepa, Panauti, and Panchkhal) representing the geographic diversity of the district. In Dolakha, we randomly selected nine public schools from a sampling frame of 337 public schools across the district using a random sampling approach. Within each selected school, we aimed to enroll up to 100 students using simple random sampling from class rosters. In Dolakha, where schools were smaller, all enrolled students were eligible to participate.

Eligibility criteria included children aged 4-18 years who were enrolled in the selected schools with no exclusion criteria besides age and school enrollment.

### Sample Size Calculation

Sample size was determined using simulation-based methods. We simulated cross-sectional serosurveys with varying sample sizes and age distributions, assuming seroincidence rates of [5-15] per 1000 person-years based on prior studies. For each scenario, we generated replicate datasets incorporating antibody kinetics parameters from confirmed cases and assay measurement error, then estimated seroincidence using maximum likelihood methods. Simulations indicated that approximately children would provide 95% confidence intervals with half-widths <[1.5] per 100 person-years, adequate for estimating seroincidence and detecting geographic differences. We targeted ∼400 children per district to balance precision with logistical feasibility.

### Data and Sample Collection

Data was collected between November 2021 and April 2022 Schools were in regular session during the study period, with no major disruptions due to holidays or COVID- 19 closures.

For each participant, we collected the following variables: age (in years), sex, grade level, school type (public/private), student grades, district of residence, home address. Age was cross-referenced with the school records, parental report and participant confirmation.

Approximately 60 µL of capillary blood was collected from each participant via finger prick onto TropBio™ filter papers (Cellabs, Brookvale, NSW, Australia). Each filter paper was labeled with a unique barcode linked to participant demographic data in a secure database. Filter papers were air-dried for at least 2 hours at room temperature, individually sealed in plastic resealable bags with desiccant, transported to Center for Infectious Disease Research and Surveillance, Dhulikhel Hospital on ice within 3-6 hours, and stored at -20°C until processing. All samples met the required criteria for processing; therefore, no samples were rejected.

### Laboratory methods

Dried blood samples were eluted by cutting single lobe of blood-filled filter paper (approximately 6 mm diameter, containing 10 µL of dried blood) by sterile scissors and submerging a filter paper protrusion in 66.5 µL of 1× PBS containing 0.05% Tween-20 buffer and incubating overnight at 4°C. Tubes were centrifuged at 10,000×g for 10 minutes and the supernatant was aliquoted(24).

Eluates were then tested for IgG responses to *Orientia tsutsugamushi*-derived recombinant 56-kDa antigen using the Scrub Typhus Detect™ ELISA kit (InBios International, Inc., Seattle, WA, USA) according to the manufacturer’s instructions. Optical density (OD) was measured at 450nm using a Bio-Rad ELISA plate reader (iMark Microplate Absorbance Reader).

All samples were tested between 27 March 2024 and 9 April 2024. Laboratory personnel were blinded to participant demographic and clinical information.

### Statistical methods

All analyses were conducted in R version 4.3.3 (R Foundation for Statistical Computing, Vienna, Austria).

We determined the seropositivity cutoff by comparing the kit-manufacturer’s recommended cutoff with a data-driven cutoff derived from using finite gaussian mixture models fit to the observed antibody response distribution. We fit 2-component Gaussian mixture models using the mixtools package (version 2.0.0) and defined the cutoff as the mean plus two and half standard deviations of the seronegative (lower) distribution(20)

IgG seroprevalence was calculated as the proportion of individuals with antibody levels above the cutoff, with 95% confidence intervals (CIs) calculated using [Wilson score method/exact binomial method]. To evaluate age-related changes in seropositivity, we fit generalized additive models (GAMs) using the *mgcv* package (version 1.9.1) with a cubic regression spline for age (25). Simultaneous 95% confidence intervals were generated using parametric bootstrap of the variance-covariance matrix (10,000 iterations) (24).

We estimated seroincidence using previously published methods that leverage antibody kinetics data from confirmed scrub typhus cases(17). In brief, this approach models the peak, time to peak, decay rate, and shape of antibody responses after infection, then uses maximum likelihood estimation to determine the most likely seroincidence rate given observed cross-sectional antibody levels, accounting for measurement error and assay detection limits(26)(27). We applied antibody decay parameters that were previously estimated from post-infection cohorts in India and Thailand(citation)(17). We used cluster-robust standard errors to account for the school-based sampling frame. The analysis was implemented in the *serocalculator* package (version 1.4.0) in R (28).

We conducted sensitivity analyses by recalculating seroprevalence using the manufacturer’s recommended cutoff ([0.37]) instead of the mixture model-derived cutoff ([0.42]).

School GPS coordinates were collected and seroprevalence/seroincidence data were mapped using the leaflet package (version 2.2.2) in R.

### Ethical procedures

The parent study received ethical approval from the Nepal Health Research Council (NHRC approval number 278/2021, dated 11 October 2021) and the Institutional Review Committee, additional supplemental approval for the scrub typhus testing was granted by Kathmandu University School of Medical Sciences (KUSMS-IRC approval number 129/24, dated 4 April 2024).

All participants (or their parents/guardians for those under 18 years) provided written informed consent prior to enrollment. For children aged 15-17, written assent was obtained in addition to parental consent. Consent forms were provided in Nepali and explained by trained study staff.

Participant data were anonymized using unique barcode identifiers, and all personal information was stored securely in password-protected databases accessible only to authorized study personnel. Participants received stationery as a gift in recognition for their participation. No serious adverse events occurred during blood collection.

## RESULTS

### Participant Characteristics

He approached 13 schools across Kavrepalanchok and Dolakha districts, of which all 13 (100%) agreed to participate (4 in Kavrepalanchok, 9 in Dolakha). Within participating schools, 1557 children were eligible based on age criteria (4-18 years), of whom 827 (53.11%) were enrolled and provided blood samples. All 827 participants had complete serological and metadata were included in the analysis. No participants were excluded, as all samples had sufficient blood volume and successfully passed serological testing. Therefore, complete data from all 827 participants were included in the final analysis (Figure 1).

**Figure 1.**
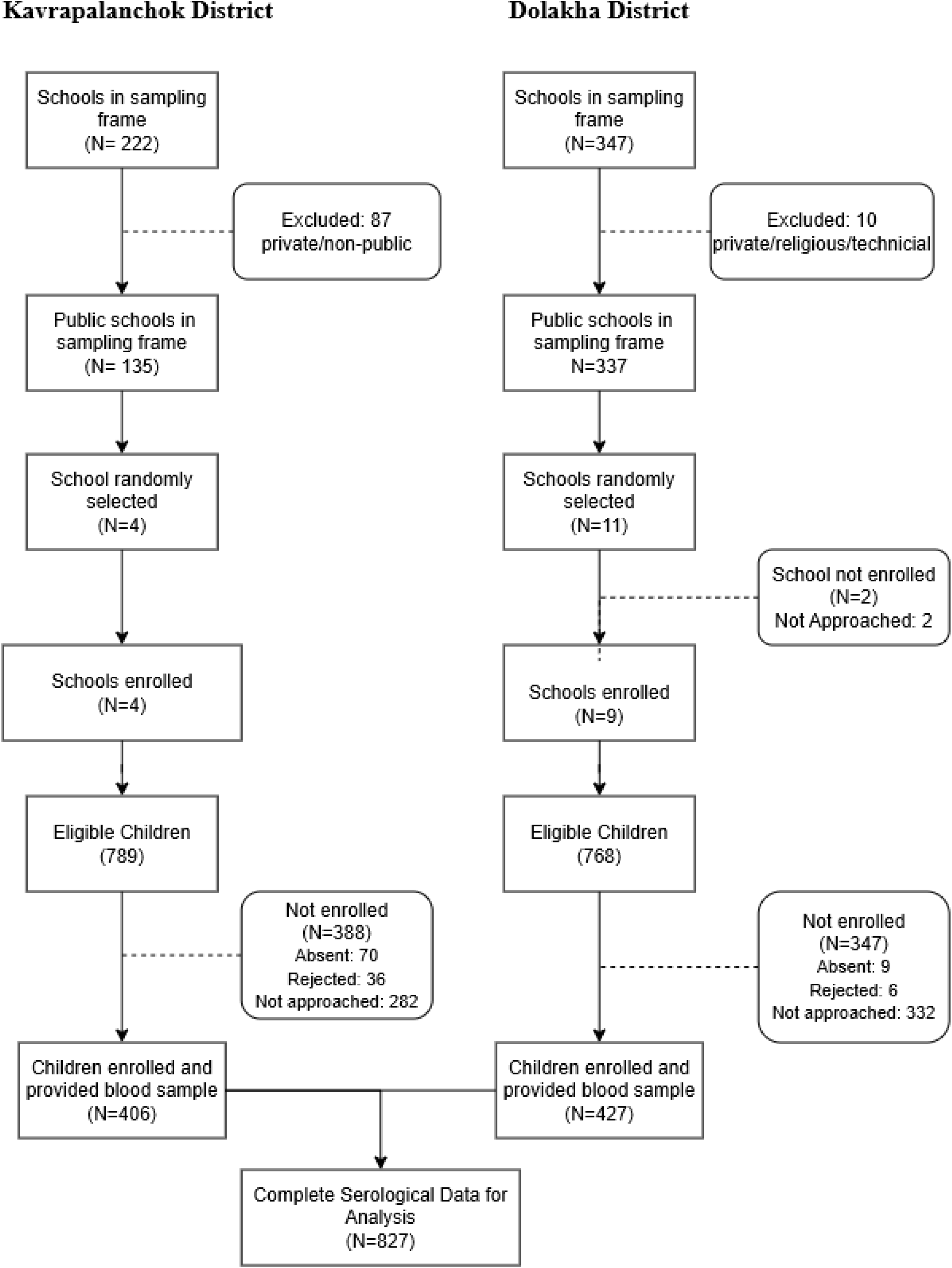
Flow Diagram.

The median age of participants was 10 years (IQR: 8-13 years). The study population included 439 females (53.1%) and 388 males (46.9%). By district, 421 participants (50.9%) were from Dolakha and 406 (49.1%) from Kavrepalanchok. Within Kavrepalanchok, participants were distributed across four municipalities: Dhulikhel (n=108, 26.6%), Banepa (n=100, 24.5%), Panauti (n=97, 23.9%), and Panchkhal (n=101, 24.9%). Within Dolakha, participants were distributed across six municipalities: Bhimeshwor(n= 62, 14.7 %), Tamakoshi (n=123, 29.2 %), Sailung (n=20, 4.8 %), and Gaurishankar (n=25, 5.0 %), Bigu (n=124 ,29.8 %), Baiteshwor (n=67, 15.9%). Based on the Degree of Urbanization (DEGURBA) in Nepal, the majority of participants attended schools in rural areas (n=522, 63.1%) compared with urban (n= 208, 25.2%), and peri-urban areas (n=97, 11.7%) (Table 1)(29).

**Table 1.**
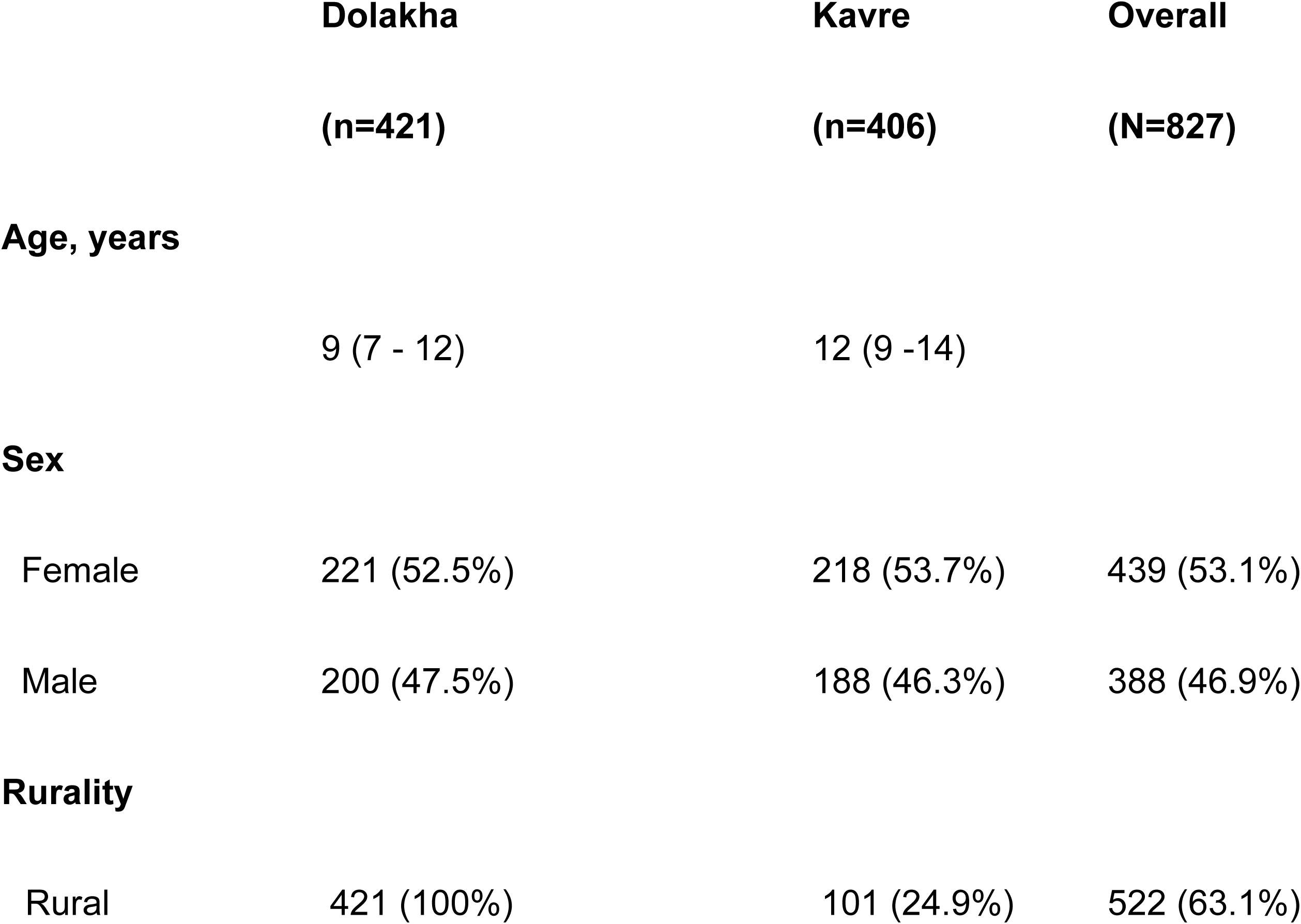
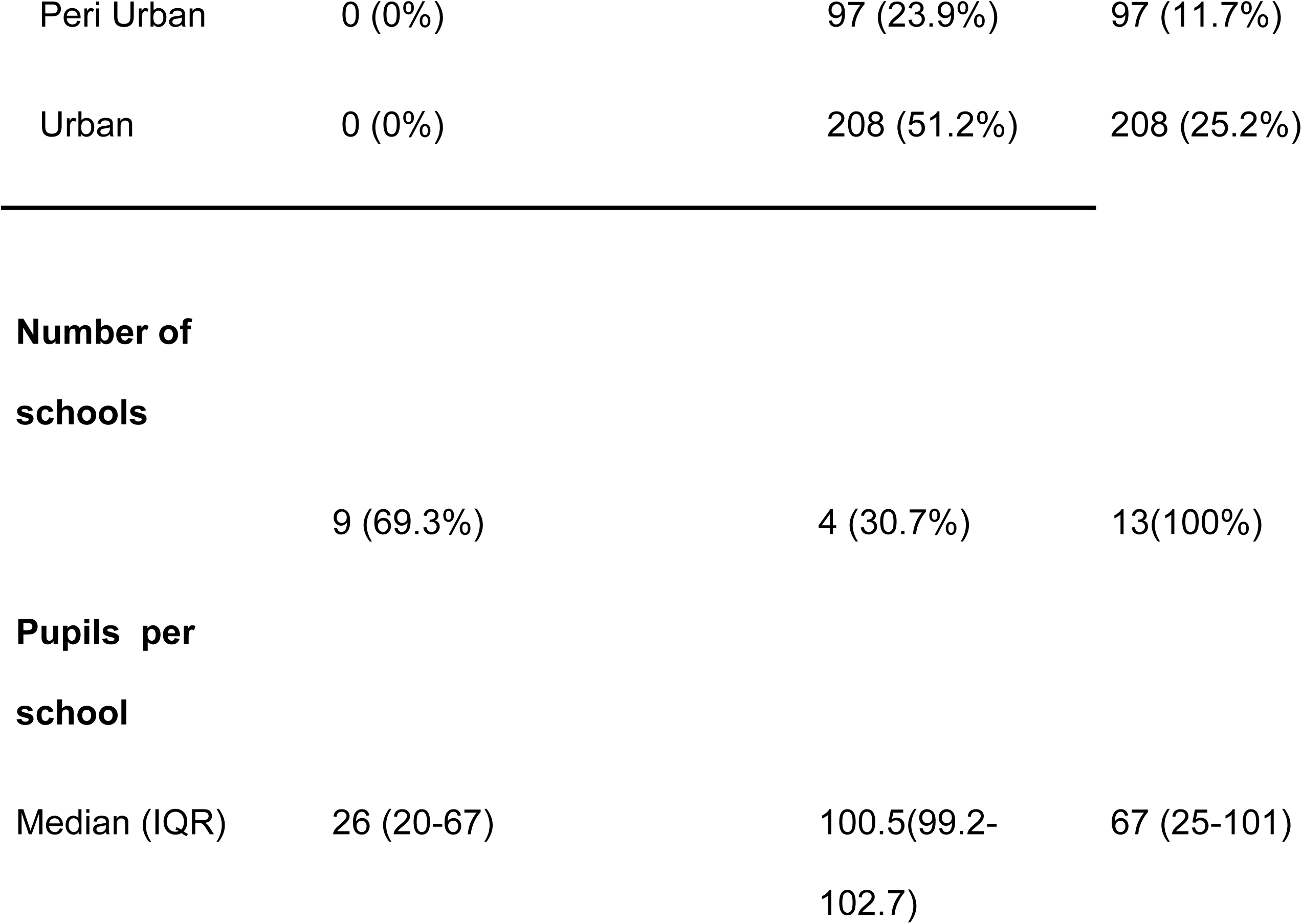
Demographic Characteristics of enrolled participants.

### Quantitative antibody responses and cutoff determination

The distribution of ELISA OD values was bimodal, with a predominant low-response population and a smaller high-response population (Figure 2A). The mixture model– derived cutoff was similar to the manufacturer-recommended cutoff, 0.42 versus 0.37, respectively, and closely aligned with the inflection point observed in the rank-order plot of OD values (Figure 2A–C). Plate-level positive and negative controls showed the expected separation, supporting assay performance across plates (Figure 2C).

**Figure 2.**
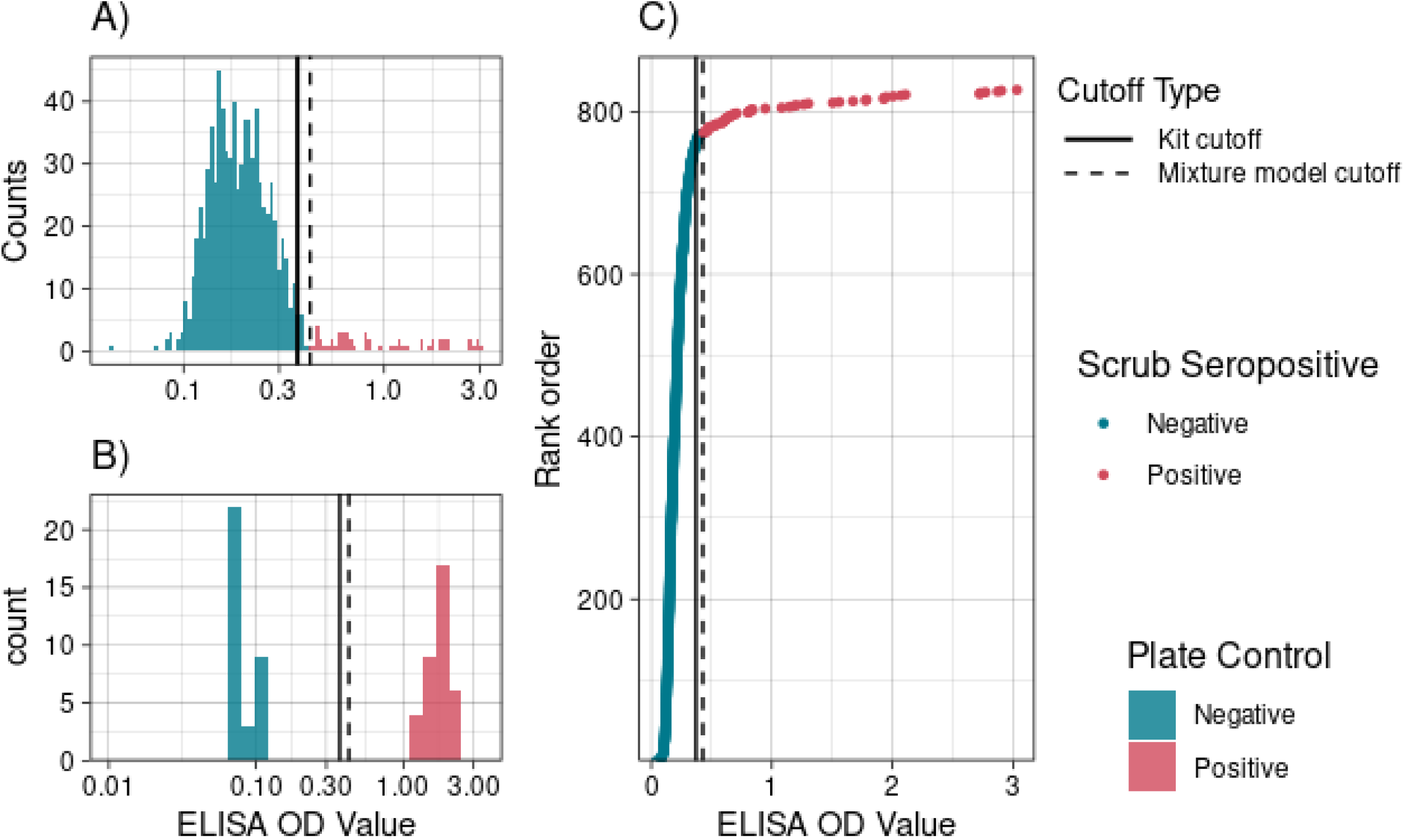
Cutoff identification methods. **A)** Histogram of ELISA optical density (OD) values from the entire study population (N = 827). **B)** Histogram of ELISA OD values for positive controls (red) and negative controls (blue) across all assay plates. **C)** Ranked OD values from the cross-sectional study population (N = 827), with each point representing an individual. Vertical lines indicate two cutoffs: The dashed line cutoff one derived from a mixture model fitted to the log-transformed OD values (mean of the lower component plus 2.5 standard deviations), and the solid lineother based on the manufacturer’s kit instructions.

### Scrub Typhus Seroincidence

Overall seroincidence was 6.3 per 1,000 person-years (95% CI: 4.7- 8.4). Seroincidence was higher in females (8.3 per 1,000 person-years; 95% CI: 5.9 -11.8) than in males (4.1 per 1,000 person-years; 95% CI: 2.4 - 6.9) (p=0.02). By district, seroincidence was 7.2 per 1,000 person-years (95% CI: 4.9 - 10.4) in Kavrepalanchok and 5.3 per 1,000 person-years (95% CI: 3.4 - 8.5) in Dolakha (p=0.32).

Seroincidence increased with age, from 3.6 per 1,000 person-years (95% CI: 1.4- 9.7) among children aged 4- 7 years to 4.0 per 1,000 person-years (95% CI: 1.9-8.4) among those aged 8- 10 years, 6.2 per 1,000 person-years (95% CI: 3.6- 10.7) among those aged 11-13 years, and 9.4 per 1,000 person-years (95% CI: 6.2-14.3) among those aged 14-18 years (Table 2, Fig 2C).

**Table 2:** Scrub typhus seroprevalence and seroincidence by district, sex, age category, and municipality.

| Category | Total tested | Number seropositive | Seroprevalence (%) (95% CI) | Seroincidence rate (per 1000-person-years) (95% CI) |
| --- | --- | --- | --- | --- |
| Overall | 827 | 54 | 6.5 (4.9–8.4) | 6.3 (95% CI: 3.4–11.6 ) |
| Dolakha | 421 | 24 | 5.7 (3.7–8.4) | 5.3 (95% CI: 2.6–11 ) |
| Kavre | 406 | 30 | 7.4 (5–10.4) | 7.2 (95% CI: 2.9–17.5 ) |
| **Gender** |  |  |  |  |
| Female | 439 | 34 | 7.7 (5.4–10.7) | 8.3 (95% CI: 4.4–15.7 ) |
| Male | 388 | 20 | 5.2 (3.2–7.8) | 4.1 (95% CI: 2.2–7.6 ) |
| **Age group** |  |  |  |  |
| 4-7 | 201 | 6 | 3 (1.1–6.4) | 3.6 (95% CI: 1.1–11.7 ) |
| 8-10 | 217 | 8 | 3.7 (1.6–7.1) | 4 (95% CI: 1.9–8.4 ) |
| 11-13 | 205 | 13 | 6.3 (3.4–10.6) | 6.2 (95% CI: 3.2–12.2 ) |
| 14-18 | 204 | 27 | 13.2 (8.9–18.7) | 9.4 (95% CI: 5–17.9 ) |
| **Municipalities** |  |  |  |  |
| Baiteshwor | 67 | 2 | 3 (0.4–10.4) | 1.7 (95% CI: 1.7–1.7 ) |
| Banepa | 100 | 8 | 8 (3.5–15.2) | 6 (95% CI: 6–6 ) |
| Bhimeshwor | 62 | 0 | 0 (0–5.8) | 0 (95% CI: 0–0 ) |
| Bigu | 124 | 6 | 4.8 (1.8–10.2) | 4.9 (95% CI: 3.8–6.3 ) |
| Dhulikhel | 108 | 17 | 15.7 (9.4–24) | 18.7 (95% CI: 18.7–18.7 ) |
| Gaurishankar | 25 | 1 | 4 (0.1–20.4) | 5.4 (95% CI: 5.4-5.4) |
| Panauti | 97 | 4 | 4.1 (1.1–10.2) | 4.3 (95% CI: 4.3–4.3 ) |
| Panchkhal | 101 | 1 | 1 (0–5.4) | 0.9 (95% CI: 0.9–0.9 ) |
| Sailung | 20 | 0 | 0 (0–16.8) | 0 (95% CI: 95% CI: 0–0) |
| Tamakoshi | 123 | 15 | 12.2 (7–19.3) | 10.3 (95% CI: 7.6–14.1 ) |

At the municipality level, seroincidence was highest in Dhulikhel (18.7 per 1,000 person-years; 95% CI: 11.6–30.2) and Tamakoshi (12.2 per 1,000 person-years; 95% CI: 5.7–14.1) respectively in Kavre and Dolakha District. Seroincidence was lowest in Panchkhal (1 per 1,000 person-years; 95% CI: 0.1- 6.7) and Baiteshwor (1.7 per 1,000 person-years; 95% CI: 0.2-12.3) (Table 2).

### Scrub Typhus Seroprevalence

Overall IgG seroprevalence was 6.5% (95% CI: 4.9-8.4%; 54/827). Seroprevalence was slightly higher in females, 7.7% (95% CI: 5.4-10.7%; 34/439), versus 5.2% (95% CI: 3.2-7.8%; 20/388) in males but the difference was not statistically significant (p=0.17).

At the district level, seroprevalence was 7.4% (95% CI: 5.0-10.4%; 30/406) in Kavrepalanchok and 5.7% (95% CI: 3.7-8.4%; 24/421) in Dolakha (p=0.40). Within Kavrepalanchok, municipality-level seroprevalence ranged from 1% (95% CI: 0-5.4%; 1/101) in Panchkhal to 15.7% (95% CI: 9.4-24%; 17/108) in Dhulikhel. Within Dolakha, seroprevalence ranged from [0] % (95% CI: 0-5.8%; 0/62) in Bhimeshwor to 12.2% (95% CI: 7-19.3%; 15/123) in Tamakoshi (Table 2, Figure 3A).

**Figure 3:**
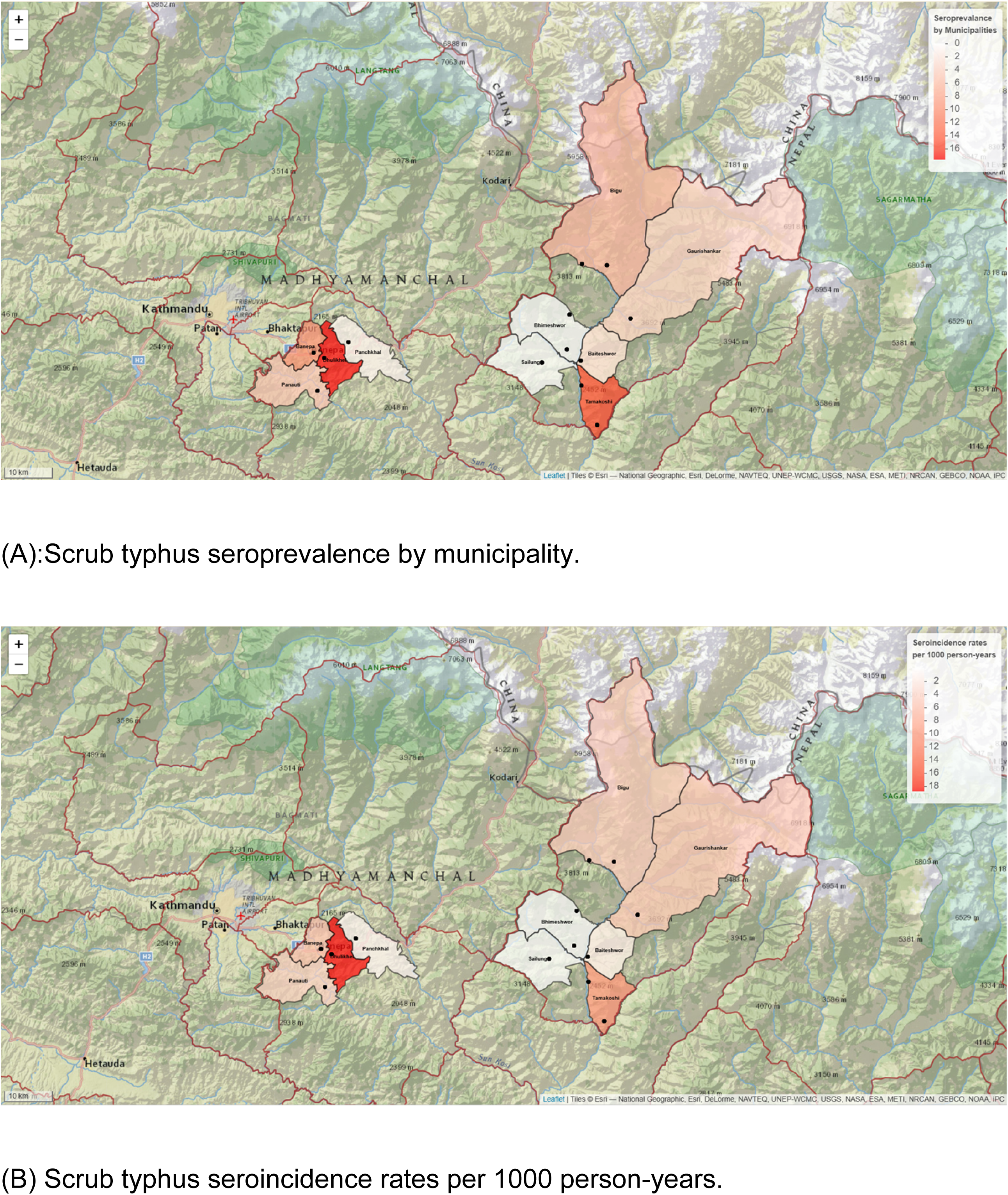
Geographic distribution of scrub typhus seroprevalence and seroincidence. (A) Seroprevalence (%) and (B) seroincidence (per 1,000 person-years) by municipality in Kavrepalanchok and Dolakha districts, Nepal, November 2021-April 2022. Black dots indicate locations of enrolled schools. Seroprevalence increased significantly with age: 3% (95% CI: [1.1-6.4] %; 6/201) among children aged 4-7 years, 3.7% (95% CI: 1.6-7.1% 8/217) among those aged 8-10 years, 6.3% (95% CI: 3.4-10.6%; 13/205) among those aged 11-13 years and 13.2% (95% CI: 8.9-18.7%; 27/204) among those aged 14-18 years (Figure 3B) (p<0.001).

**Figure 4:**
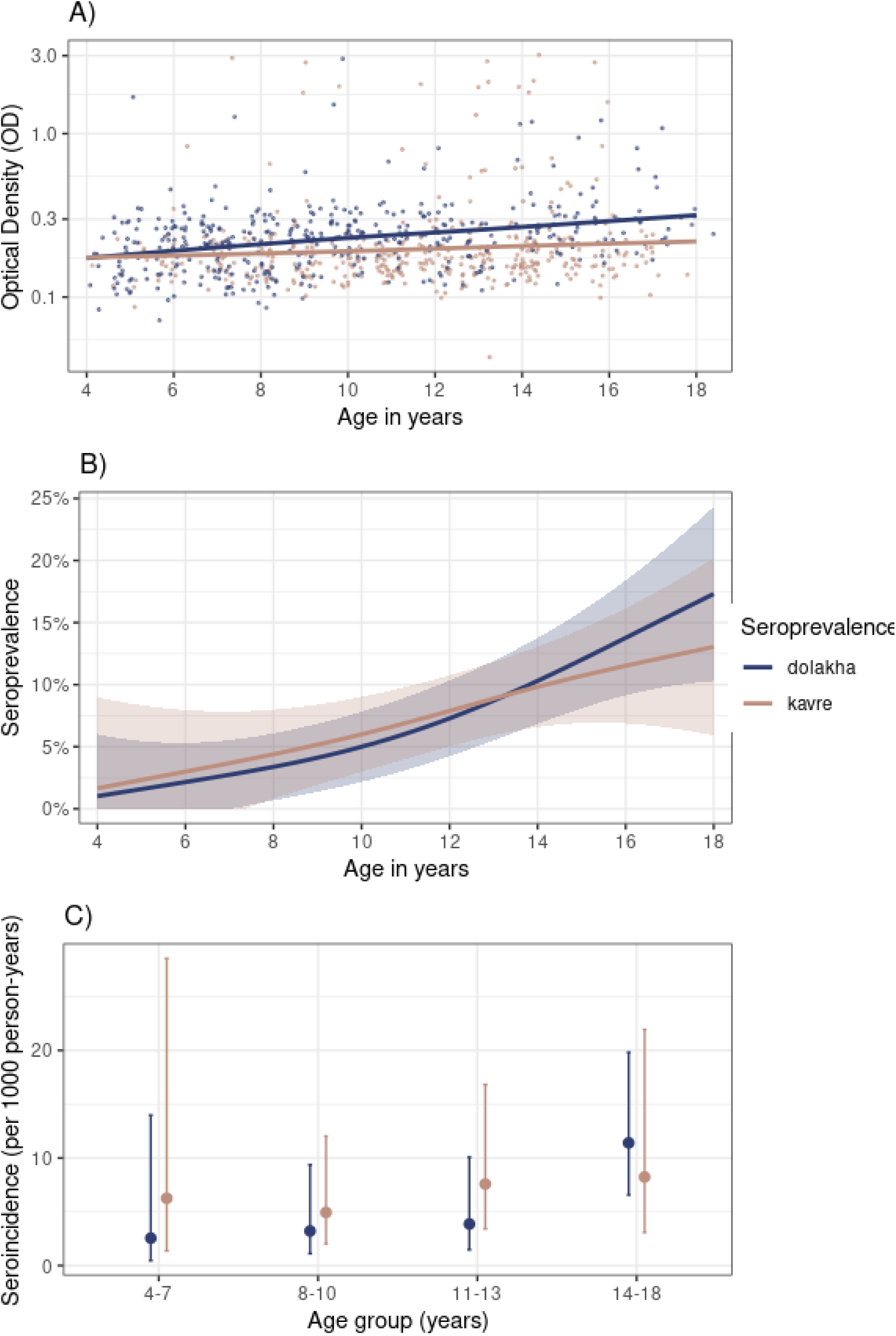
Age-related patterns in scrub typhus antibody responses, seroprevalence, and seroincidence. Figure 4: (A) Quantitative IgG antibody responses (optical density) as a function of age. Each point represents an individual participant (N=827). The horizontal dashed line indicates the seropositivity cutoff (OD=[0.42]). (B) Seroprevalence as a function of age. Points represent observed seroprevalence in [X-year] age bins; the solid line represents the fitted generalized additive model (GAM) with 95% simultaneous confidence intervals (shaded area). (C) Seroincidence as a function of age, showing relatively constant force of infection across age groups. Error bars represent 95% confidence intervals."

### Sensitivity Analyses

Using the manufacturer’s recommended cutoff ([0.37]) instead of the mixture model cutoff yielded slightly higher overall seroprevalence of 7.9% (95% CI: 6.2–10.0%; 65/827).

## Discussion

This school-based serosurvey reveals a substantial burden of scrub typhus among school-age children in Kavrepalanchok and Dolakha districts, Nepal. Seroprevalence and seroincidence showed broadly concordant patterns across districts and age groups among younger children. At older ages, however, seroprevalence and seroincidence estimates began to diverge, consistent with the accumulation of IgG with increasing cumulative exposure over time. Both seroprevalence and seroincidence were higher among females, suggesting possible sex-based differences in exposures among school-age children. Together, these patterns underscore the high and underrecognized burden of scrub typhus among children in periurban and rural Nepal.

Seroprevalence rose progressively across age groups, with a particularly steep increase in the oldest group more than doubling from the 11–13 year age group to the 14–18 year age group. In contrast, seroincidence increased only modestly across age groups, and the difference was not statistically significant. This divergence is expected: IgG responses to *O. tsutsugamushi* persist for years after infection, with levels remaining above diagnostic cutoffs for more than 36 months in most individuals(17,30). Seroprevalence therefore reflects cumulative past exposure and will increase with age simply because older individuals have had more years to be infected, even if the underlying force of infection is relatively constant. Seroincidence, by contrast, estimates the rate of new infections per unit time(17).

Both seroprevalence and seroincidence were higher among females than males, with seroincidence estimates suggesting that girls experienced approximately twice the force of infection compared with boys. This pattern is consistent with the Devamani et al. population-based cohort study in rural South India, where the incidence of clinical scrub typhus infection was higher among female participants[4]. Schmidt et al., analyzing the same South Indian cohort for risk factors, found that the increased risk in females compared with males was unaffected by adjustment for agricultural and outdoor activities, suggesting that occupational exposure alone does not explain the sex-based difference(15). The finding of a higher force of infection among girls in a school-age population observed in this study is noteworthy, as this group would not be expected to have the occupational differences — such as agricultural labor that are commonly invoked to explain higher scrub typhus rates among adult women(31). Instead, the difference may reflect sex-specific patterns in domestic chores or peridomestic activities that differentially expose girls to chigger mite habitats. In rural Nepal, girls may partake in chores such as grass cutting for animal fodder, collecting firewood, and other activities that bring them into contact with vegetation and soil(32,33). Whether and how these specific activities differ by sex among Nepalese school children and contribute to the observed difference in force of infection warrants investigation in future research.

At the district level, seroprevalence and seroincidence were modestly but non- significantly higher in Kavrepalanchok than Dolakha, with overlapping confidence intervals indicating broadly comparable transmission intensity between the two districts. Within districts, however, substantial municipality-level heterogeneity was observed: seroprevalence ranged from 0% in Bhimeshwor and Sailung to 16% in Dhulikhel, with seroincidence following a concordant pattern. This focal geographic variation likely reflects local ecological factors that influence human–vector contact, and underscores that district-level estimates may mask important local variation in transmission risk.

Both districts are predominantly agricultural and lie within Nepal’s monsoon zone, though they differ in topography and rainfall. Dolakha is a rural mountainous district with elevations ranging from 723 to 7,134 meters and annual rainfall of 1,500–2,000 mm, while Kavrepalanchok has a more moderate landscape (280–3,018 meters elevation) with annual rainfall of 1,200–1,500 mm. Prior studies have identified agricultural occupations as a key risk factor for scrub typhus, as humans are more likely to encounter chigger mites when working or walking through fields and grasslands(34). Higher rainfall and greater vegetation density may support larger populations of chigger mites and their rodent hosts, potentially contributing to focal areas of high transmission within both districts. Environmental suitability modeling in Nepal has shown that areas close to agricultural land with gentle slopes have higher suitability for scrub typhus occurrence, with approximately 43% of the population living in highly suitable areas(35).

This school-based serosurvey was conducted over a period of six months and required four research staff. Participation rates were high in both districts with 91.8 and 98.5 in Kavrepalanchok and Dolakha respectively. Importantly, the seroincidence estimates obtained from this school-based approach were comparable to those derived from a population-based serosurvey of 0- to 27-year-olds in the Kavrepalanchok district, which estimated a seroincidence of 14 per 1,000 person-years(17). While population-based serosurveillance studies are generally less resource-intensive than clinical surveillance, school-based surveillance may offer an even more efficient approach for characterizing the burden of scrub typhus and other neglected tropical diseases in endemic settings(36). Schools provide a readily accessible sampling frame with high participation rates, and the age range of school children (4-18 years) captures a period of increasing cumulative exposure that is informative for estimating transmission intensity.

Several limitations should be considered when interpreting these results. First, the study was conducted among school-age children, and findings cannot be extrapolated to younger children not yet enrolled in school or to adult populations, who may have different exposure patterns and cumulative seropositivity. Second, only children enrolled in public schools were included; children attending private schools may have different socioeconomic and environmental exposures, which could affect generalizability. Finally, the study was conducted between November and April, which spans the post-monsoon and dry seasons.Scrub typhus transmission in Nepal peaks during the monsoon and post-monsoon (August - October) and the timing of sample collection may not fully capture seasonal variation in recent infections and may have introduced bias. However, IgG seroprevalence reflects cumulative rather than acute exposure and is therefore less sensitive to sampling season(9,37).

## Conclusion

This study reveals a substantial burden of scrub typhus among school-age children in Kavrepalanchok and Dolakha districts, Nepal. Seroprevalence and seroincidence provided complementary insights: seroprevalence captured the expected age-dependent accumulation of past exposure, while seroincidence offered a more stable estimate of recent transmission intensity across age groups. Both measures indicated a higher force of infection among girls, suggesting that sex-specific domestic or peridomestic activities may contribute to differential exposure even among school-age children. Substantial municipality-level heterogeneity in both measures highlights the focal nature of scrub typhus transmission. These findings demonstrate that school-based serosurveys paired with seroincidence estimation are an efficient and informative approach for characterizing scrub typhus transmission intensity at the population level. Together, these results confirm that scrub typhus is an important and underrecognized cause of pediatric infectious disease exposure in Nepal and support the integration of serosurveillance into neglected tropical disease control programs in endemic settings.

## Data Availability

De-identified data will be available upon publication in Open Science Framework (OSF)

## Acknowledgment

We would like to acknowledge the essential contributions of field team Nisha Shrestha, Manisha Banjara, Anil Khanal, Neeru Suwal. We are grateful to all the participating schools and participants (students) for their valuable time and responses. We thank InBios International Inc. for providing Scrub Typhus Detect™ ELISA kits used in this study. The company had no role in study design, data collection, analysis, interpretation, or manuscript preparation.

## Funding

This study was supported by a grant from the Gates Foundation (INV-000572). K.A. was additionally supported by the Fogarty International Center of the National Institute of Health (award no. K01TW012177) and National Institute of Allergy and Infectious Diseases of the National Institutes of Health (R21AI176416). The funders had no role in study design, data collection, data analysis, data interpretation, or writing of the manuscript.

## Notes

### Competing Interest Statement

The authors have declared no competing interest.

